# An umbrella review of the facilitators and barriers to implementing Artificial Intelligence (AI) solutions within hospital settings: through the lens of the NASSS framework (spread, scale-up and sustainability)

**DOI:** 10.1101/2025.06.19.25329916

**Authors:** Sigrún Eyrúnardóttir Clark, Sanjoli Mathur, Yolanda Barrado-Martín, Ahmed El-Sayed, Yue Zhao, Zarnie Khadjesari, Fiona Stevenson, Laurence Lovat, Cecilia Vindrola-Padros

## Abstract

Advancements in artificial intelligence (AI) are revolutionising the healthcare sector, but challenges exist in AI adoption and its long-term use. This umbrella review aimed to identify the facilitators and barriers of AI implementation within hospitals and was registered on PROSPERO. Five databases (MEDLINE, HMIC, CINAHL Plus, Web of Science and Cochrane Reviews) were searched in January 2025, 763 articles were screened, with 13 included. The inclusion criteria encompassed studies implementing AI that were conducted within the hospital setting. The quality of the data were assessed using the ROBIS checklist and data were extracted using the NASSS (Nonadoption, Abandonment, and challenges to the Scale-up, Spread, and Sustainability) framework, to demonstrate how AI implementation was affected by: whether the AI solution had been technologically validated to ensure generalisability across departments; evidence the AI solution brings measurable gains; a lack of trust or understanding among hospital staff; the budgets and resources available to onboard the AI solution, train staff, and maintain the solution; the need for national policies on funding and regulating AI solutions. These factors affected the adoption, spread, scalability and sustainability of AI implementation and could be considered in future implementation efforts. The study was funded by the NIHR (NIHR205439).

## Introduction

Advancements in artificial intelligence (AI) are revolutionising the healthcare sector^1,2^. AI offers potential economic advantages, enhances medical services, and optimises the use of resources within hospitals^1,2^. AI is an overarching system that processes large bodies of data and is able to learn from the data to solve problems through a subset of analytical techniques such as machine learning^2,3^. Machine learning is the process in which a computer system is able to identify patterns, learn from those patterns, and then perform actions to produce results^3^. A machine learning algorithm will automatically adapt its algorithm based on its experience of receiving repetitions of sample data along with desired outcomes, this is a process known as training the machine learning algorithm. As a result, the algorithm will produce the desired outcome from the training sample data and should also be able to generalise its algorithm to produce the desired outcome from new (non-sample) data^4^. Despite AI’s transformative potential in healthcare, its adoption and long-term use is shaped by various factors, including education, organisational and financial factors within institutions, and regulatory policies^5^. Identifying the factors that could act as barriers in implementation can help facilitate the implementation of AI solutions across healthcare systems.

Existing research, including systematic reviews by Hassan et al. and Ahmed et al., has examined an array of barriers and facilitators to AI implementation in healthcare^2,6^. Key hindering factors identified by these publications included insufficient IT infrastructure, high upfront costs, and ambiguous legal frameworks surrounding liability. Conversely, influential facilitators such as transparent governance structures, well-defined protocols for data security, and ongoing professional development have been shown to bolster acceptance and adoption rates. There have also been numerous systematic reviews focussing on the barriers and facilitators of AI implementation within specific healthcare contexts such as acute care^7^, nursing^8^ and infection control^9^. Although these reviews highlight critical insights into how AI integration into healthcare workflows is influenced by various factors in specific clinical areas, no umbrella review has been conducted to explore cross-cutting factors across clinical settings and types of AI solutions. The aim of this umbrella review is to identify common trends in implementation and collate the factors acting as barriers and facilitators to implementing AI in hospital settings. Addressing this gap would enable the field to glean a more cohesive understanding of the multifaceted issues at play and develop tailored strategies for embedding AI solutions more seamlessly into hospital settings.

## Methods

This umbrella review was developed in accordance with the Preferred Reporting Items for Overviews of Reviews (PRIOR) guidelines outlined by Gates et al. (2022)^10^. The protocol for the review was published on PROSPERO (CRD 42025638087)^11^.

### 1. Search strategy

A scoping search of the existing literature was conducted on Google Scholar. As a result of this process the combination of the following search terms were employed: ‘Artificial Intelligence’ AND ‘Hospital’ AND ‘Systematic review’ across five databases: MEDLINE, CINAHL Plus, HMIC, Cochrane Reviews and Web of Science in January 2025. The detailed search criteria can be found in Appendix 1.

### 2. Inclusion and exclusion criteria

The eligibility criteria that were used for the review can be found in Table 1. The types of studies that could be included encompassed systematic reviews, literature reviews, scoping reviews/rapid reviews. There were no limitations on the types of study design that could be included, so quantitative, qualitative and mixed method study designs were all relevant. Hospital settings were the only type of clinical setting that would be included in the review, and AI and its subset technologies were considered. There were no exclusions based on the country of the study. Publications were limited to those that shared the perspectives on the barriers and facilitators to implementing AI in the hospital setting. Studies not mentioning AI, those conducted outside hospital environments (e.g., dental practices, primary care clinics), or articles that were single empirical studies rather than reviews were excluded.

**Table 1.** Inclusion criteria.

|  |  |
| --- | --- |
| **Inclusion** | **Exclusion** |
| Use of artificial intelligence (AI) or any subset techniques.<br>• AI as an overarching system that processes large bodies of data through a subset of analytical techniques.<br>• Subset of analytical techniques such as machine learning, natural language processing, deep learning etc. | No mention of AI or subset techniques |
| Services, interventions or approaches conducted in the hospital setting. | Not a hospital setting (e.g. dentist or GP). |
| Systematic review, meta-synthesis, meta-analysis, literature review, scoping review. | Not a review of publications (e.g. single empirical study). |
| Implementation of AI; or barriers or facilitators to AI implementation. |  |

### 3. Study selection

Once searches were retrieved from the scientific databases, the results were de-duplicated using Endnote and were then imported into the software Rayyan^12^ for title and abstract screening. Four researchers conducted screening of the same 25% of title and abstracts and underwent team discussions to identify discrepancies and reconfirm the teams understanding of the eligibility criteria. Two researchers then independently screened the remaining publications. A similar process was then initiated with full text screening, with the group of researchers conducting screening of the same 25% of full texts, followed by team discussions to identify discrepancies and build confidence for two researchers to then screen the remaining full texts.

### 4. Data extraction

A data extraction form was developed using Microsoft Excel based on the Nonadoption, Abandonment, Scale-Up, Spread, and Sustainability (NASSS) framework^13^. The data extraction form was piloted among the research team who extracted the same 25% of included publications to build confidence and ensure alignment, three researchers then independently extracted data from the remaining publications.

### 5. Critical appraisal

The ROBIS tool is a critical appraisal tool for assessing the risk of bias in systematic reviews^14^. ROBIS was utilised to evaluate literature quality included in the review across different study designs which included qualitative, quantitative, and mixed methods studies. The quality assessment was conducted by three researchers, one conducting the initial quality appraisal, and two additional researchers cross-checking half of the decisions each.

### 6. Synthesis

The content from the data extraction form was synthesised using narrative synthesis to deductively group together content that could fit under the categories from the NASSS framework.

## Results

### 1. Study Characteristics

The search was performed across five databases, yielding 963 records. After duplicates were removed, 763 records remained for title and abstract screening. After the screening of titles and abstracts, 215 articles remained and underwent a full-text review. As a result of the comprehensive review of records sourced from database searches, 9 articles were selected for data extraction. An additional 4 articles were also selected for data extraction; these articles were handpicked from the scoping process conducted prior to implementing the umbrella review. An overview of the results from the screening process can be found in Figure 1.

**Figure 1.**
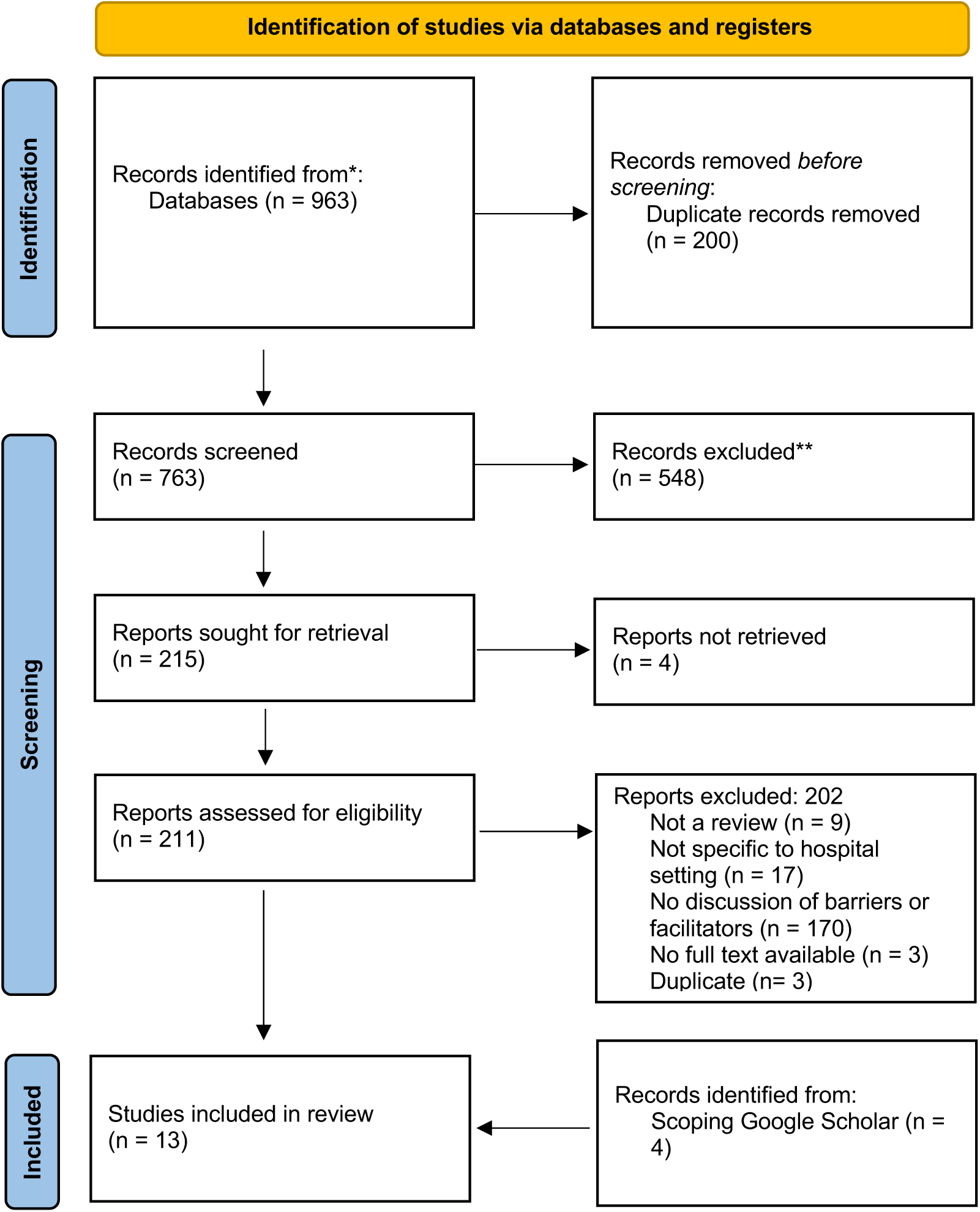
PRISMA flow diagram^15^.

All studies were ranked with a high risk of bias based on the ROBIS scores, as demonstrated in Table 2. The researchers agreed on the overall high-risk score for each publication; there were more granular discrepancies, which were then discussed between the researchers. Some of the ROBIS criteria were not applicable to the qualitative studies in the umbrella review, as the criteria were mainly relevant to quantitative meta-analyses. This may have affected the overall ROBIS scores and is a limitation of the research.

**Table 2.**
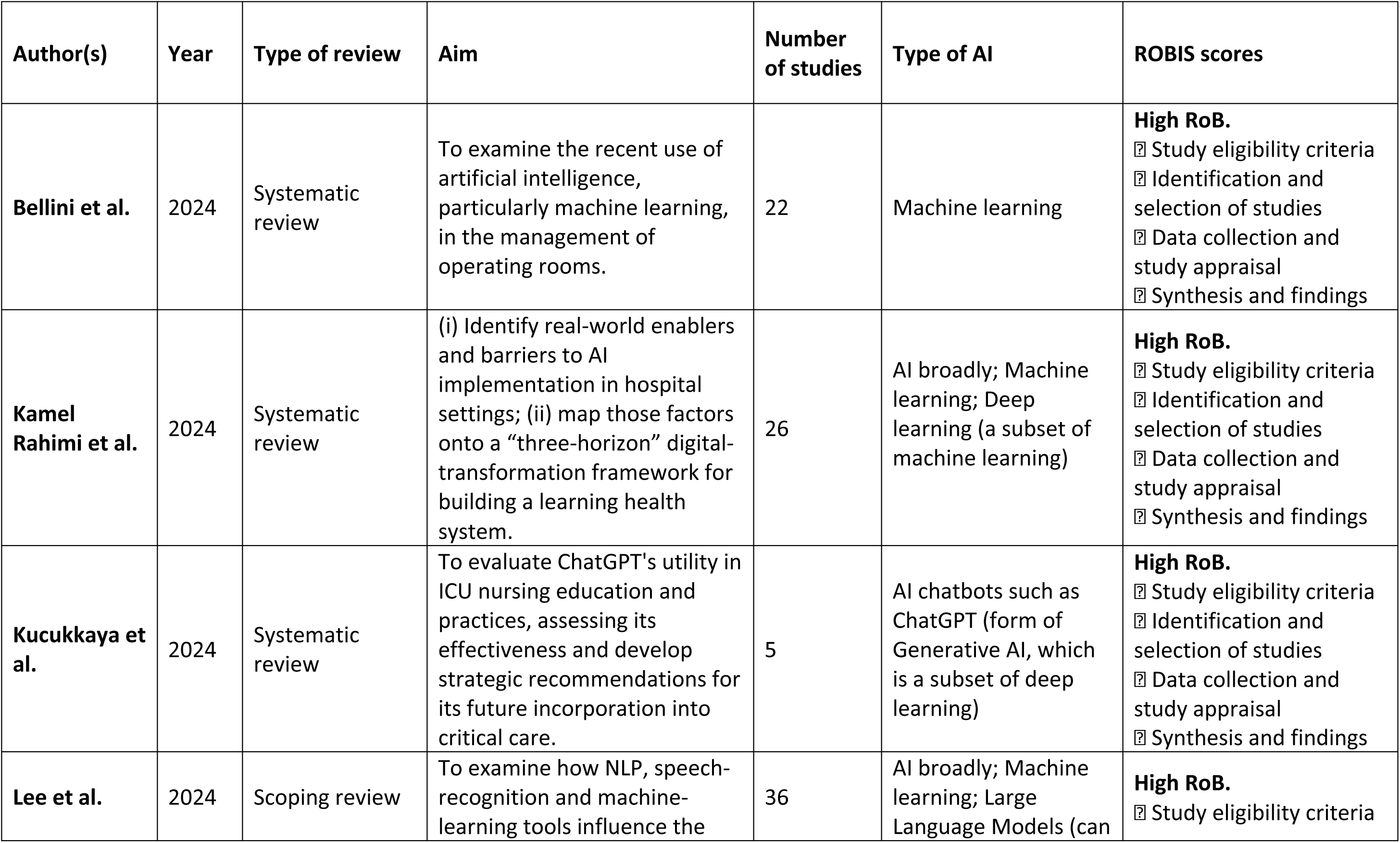
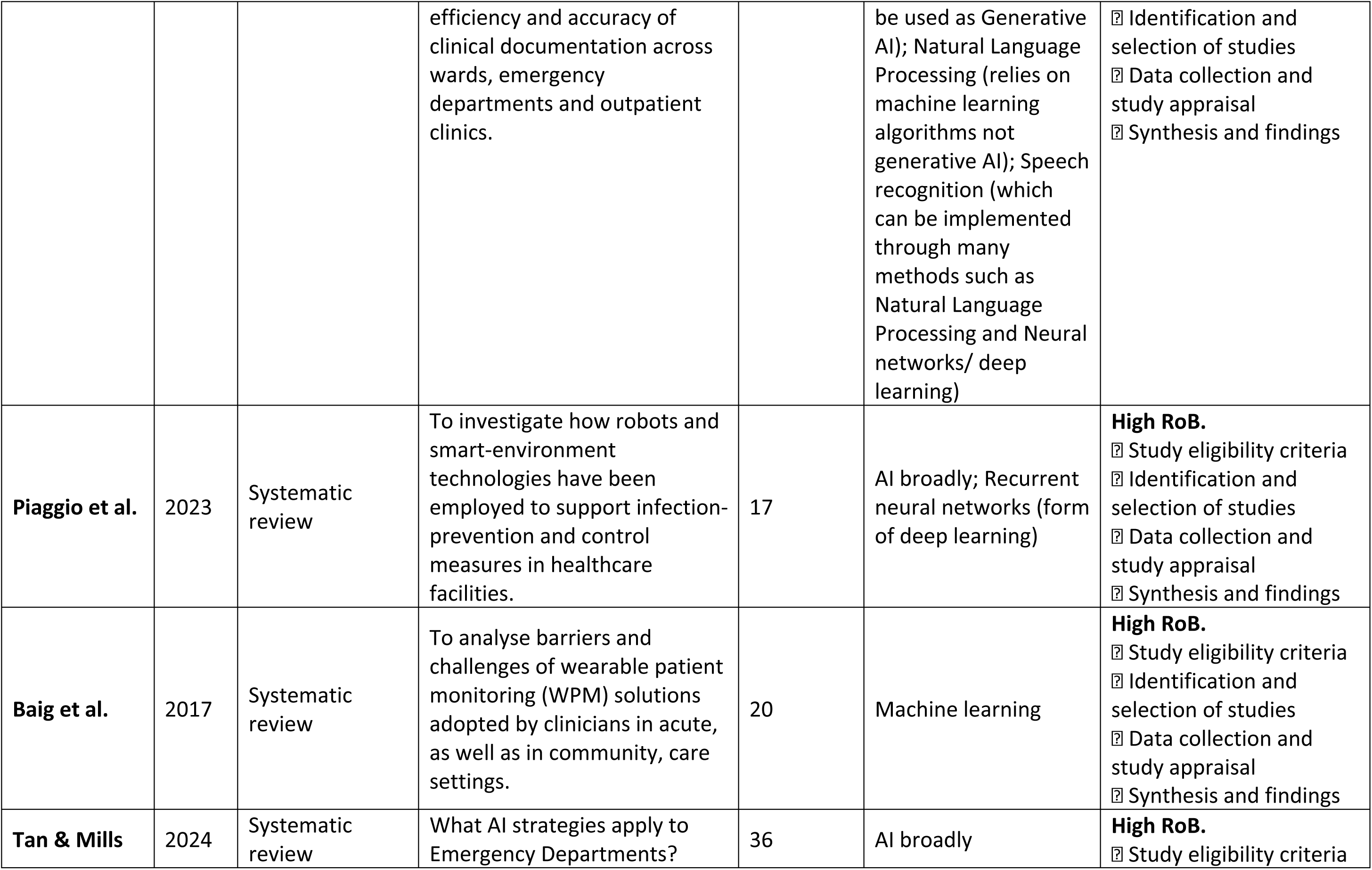
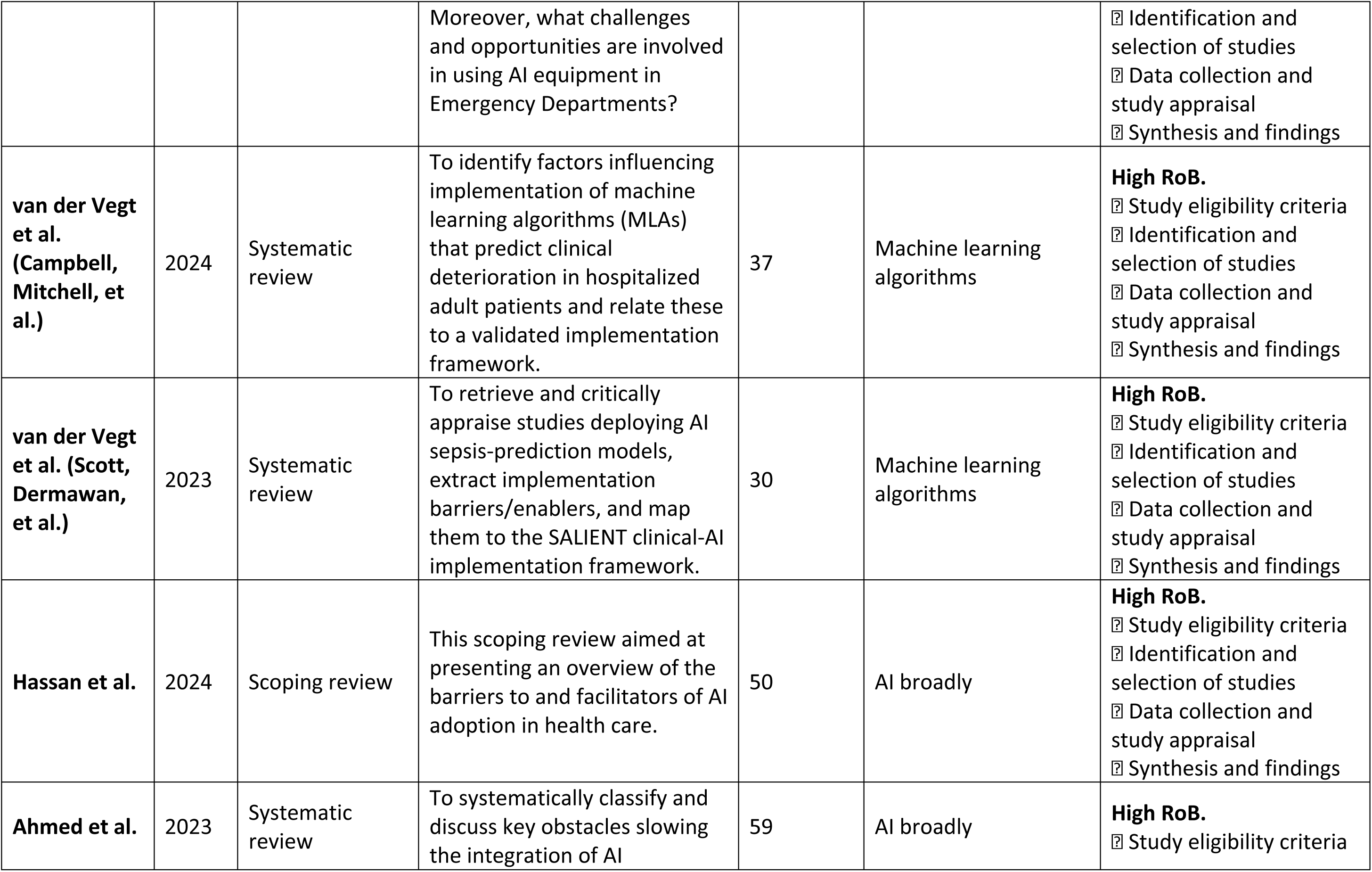
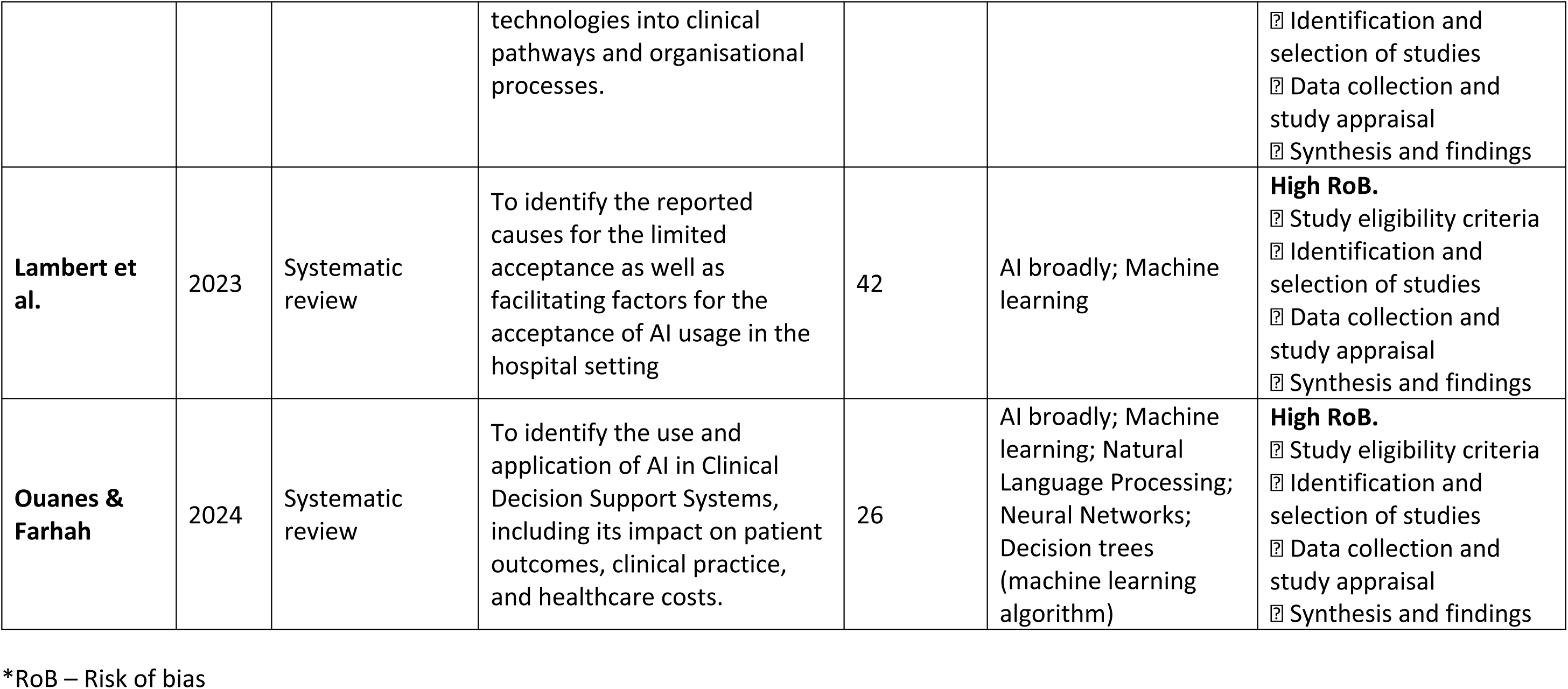
Study characteristics.

| Author(s) | Year | Type of review | Aim | Number of studies | Type of AI | ROBIS scores |
| --- | --- | --- | --- | --- | --- | --- |
| Bellini et al. | 2024 | Systematic review | To examine the recent use of artificial intelligence, particularly machine learning, in the management of operating rooms. | 22 | Machine learning | **High RoB.**<br>☐ Study eligibility criteria<br>☐ Identification and selection of studies<br>☐ Data collection and study appraisal<br>☐ Synthesis and findings |
| Kamel Rahimi et al. | 2024 | Systematic review | (i) Identify real-world enablers and barriers to AI implementation in hospital settings; (ii) map those factors onto a “three-horizon” digital-transformation framework for building a learning health system. | 26 | AI broadly; Machine learning; Deep learning (a subset of machine learning) | **High RoB.**<br>☐ Study eligibility criteria<br>☐ Identification and selection of studies<br>☐ Data collection and study appraisal<br>☐ Synthesis and findings |
| Kucukkaya et al. | 2024 | Systematic review | To evaluate ChatGPT's utility in ICU nursing education and practices, assessing its effectiveness and develop strategic recommendations for its future incorporation into critical care. | 5 | AI chatbots such as ChatGPT (form of Generative AI, which is a subset of deep learning) | **High RoB.**<br>☐ Study eligibility criteria<br>☐ Identification and selection of studies<br>☐ Data collection and study appraisal<br>☐ Synthesis and findings |
| Lee et al. | 2024 | Scoping review | To examine how NLP, speech-recognition and machine-learning tools influence the | 36 | AI broadly; Machine learning; Large Language Models (can | **High RoB.**<br>☐ Study eligibility criteria |
|  |  |  | efficiency and accuracy of clinical documentation across wards, emergency departments and outpatient clinics. |  | be used as Generative AI); Natural Language Processing (relies on machine learning algorithms not generative AI); Speech recognition (which can be implemented through many methods such as Natural Language Processing and Neural networks/ deep learning) | ☐ Identification and selection of studies<br>☐ Data collection and study appraisal<br>☐ Synthesis and findings |
| **Piaggio et al.** | 2023 | Systematic review | To investigate how robots and smart-environment technologies have been employed to support infection-prevention and control measures in healthcare facilities. | 17 | AI broadly; Recurrent neural networks (form of deep learning) | **High RoB.**<br>☐ Study eligibility criteria<br>☐ Identification and selection of studies<br>☐ Data collection and study appraisal<br>☐ Synthesis and findings |
| **Baig et al.** | 2017 | Systematic review | To analyse barriers and challenges of wearable patient monitoring (WPM) solutions adopted by clinicians in acute, as well as in community, care settings. | 20 | Machine learning | **High RoB.**<br>☐ Study eligibility criteria<br>☐ Identification and selection of studies<br>☐ Data collection and study appraisal<br>☐ Synthesis and findings |
| **Tan & Mills** | 2024 | Systematic review | What AI strategies apply to Emergency Departments? | 36 | AI broadly | **High RoB.**<br>☐ Study eligibility criteria |
|  |  |  | Moreover, what challenges and opportunities are involved in using AI equipment in Emergency Departments? |  |  | ? Identification and selection of studies<br>? Data collection and study appraisal<br>? Synthesis and findings |
| **van der Vegt et al. (Campbell, Mitchell, et al.)** | 2024 | Systematic review | To identify factors influencing implementation of machine learning algorithms (MLAs) that predict clinical deterioration in hospitalized adult patients and relate these to a validated implementation framework. | 37 | Machine learning algorithms | **High RoB.**<br>? Study eligibility criteria<br>? Identification and selection of studies<br>? Data collection and study appraisal<br>? Synthesis and findings |
| **van der Vegt et al. (Scott, Dermawan, et al.)** | 2023 | Systematic review | To retrieve and critically appraise studies deploying AI sepsis-prediction models, extract implementation barriers/enablers, and map them to the SALIENT clinical-AI implementation framework. | 30 | Machine learning algorithms | **High RoB.**<br>? Study eligibility criteria<br>? Identification and selection of studies<br>? Data collection and study appraisal<br>? Synthesis and findings |
| **Hassan et al.** | 2024 | Scoping review | This scoping review aimed at presenting an overview of the barriers to and facilitators of AI adoption in health care. | 50 | AI broadly | **High RoB.**<br>? Study eligibility criteria<br>? Identification and selection of studies<br>? Data collection and study appraisal<br>? Synthesis and findings |
| **Ahmed et al.** | 2023 | Systematic review | To systematically classify and discuss key obstacles slowing the integration of AI | 59 | AI broadly | **High RoB.**<br>? Study eligibility criteria |
|  |  |  | technologies into clinical pathways and organisational processes. |  |  | ☐ Identification and selection of studies<br>☐ Data collection and study appraisal<br>☐ Synthesis and findings |
| **Lambert et al.** | 2023 | Systematic review | To identify the reported causes for the limited acceptance as well as facilitating factors for the acceptance of AI usage in the hospital setting | 42 | AI broadly; Machine learning | **High RoB.**<br>☐ Study eligibility criteria<br>☐ Identification and selection of studies<br>☐ Data collection and study appraisal<br>☐ Synthesis and findings |
| **Ouanes & Farhah** | 2024 | Systematic review | To identify the use and application of AI in Clinical Decision Support Systems, including its impact on patient outcomes, clinical practice, and healthcare costs. | 26 | AI broadly; Machine learning; Natural Language Processing; Neural Networks; Decision trees (machine learning algorithm) | **High RoB.**<br>☐ Study eligibility criteria<br>☐ Identification and selection of studies<br>☐ Data collection and study appraisal<br>☐ Synthesis and findings |

\*RoB – Risk of bias

Most reviews focused on the use of AI or machine learning-based approaches broadly, whilst one review focused specifically on the use of generative AI chatbots in the form of ChatGPT^8^. After checking the individual studies included in each review, the research team identified that seven of the 13 reviews in our umbrella review, contained some of the same studies.

The following table summarises the 13 articles included in the umbrella review.

### 2. The reported factors acting as facilitators and barriers aligned with the NASSS framework

The Nonadoption, Abandonment, and challenges to the Scale-up, Spread, and Sustainability (NASSS) framework has been used to show how AI adoption in the hospital setting depends on (1) the condition, (2) the technology, (3) the value proposition, (4) the adopter system, (5) the organisation, (6) the wider context, and (7) how the domains interact with each other over time. Table 3 highlights these seven domains and summarises the key themes within each domain identified from the umbrella review.

**Table 3.** Summary of the themes within the NASSS domains.

| NASSS Domain | Key Factors Identified |
| --- | --- |
| **Domain 1:**<br>**The Condition** | An algorithms performance can be affected by how an outcome measure of a condition is defined |
| **Domain 2:**<br>**The Technology** | AI solution needs to be able to integrate in the existing computational resources |
|  | Concerns around the accuracy of outputs from the AI |
|  | Concerns around the quality and relevance of the data being fed into AI solutions |
|  | Concerns around the generalisability of the AI solution and its outputs |
|  | Lack of transparency of AI outputs |
| **Domain 3:**<br>**The Value Proposition** | The need for measurable gains through cost savings of clinical outcomes |
|  | The need for empirical evidence on the added value of the AI solution |
|  | AI solutions must be used in relation to an identified problem |
|  | Concerns that AI is not relevant to an individual's workflow |
| **Domain 4:**<br>**The Adopter System** | A lack of trust among end users due to misunderstanding outputs or concerns around accuracy |
|  | The need to train end users in interpreting AI outputs |
|  | Fear of job displacement or autonomy loss |
|  | The need to co-design AI solutions with end users |
|  | The need to work with multi-disciplinary teams who implement AI solutions |
| **Domain 5:**<br>**The Organisation** | Stable funding essential for implementing and maintaining AI solutions |
|  | Support from senior leadership to roll out AI solutions |
|  | Non-disruptive integration of AI into existing workflows |
|  | Formalised implementation plans for the AI solution |
| **Domain 6:**<br>**The Wider**<br>**Context** | Financial and legal endorsement from governmental or national bodies |
|  | National regulatory standards based on implementation of AI solutions |
|  | An agreed national standard for the accuracy of AI before its approved for clinical practice |
|  | Existing financial challenges within health systems |
|  | Inequalities between hospital sites can affect access to costly AI solutions |
| **Domain 7:**<br>**Interactions**<br>**& Adaptation**<br>**Over Time** | Complex outputs from AI solutions could lead to a lack of understanding among hospital staff |
|  | Ongoing staff feedback can improve the AI solution |

#### Domain 1: The Condition

The first domain of the Nonadoption, Abandonment, and challenges to the Scale-up, Spread, and Sustainability (NASSS) framework concerns the clinical characteristics of the condition for which a technology is introduced. Very few reviews in this umbrella analysis addressed the extent to which characteristics of specific conditions may impede or facilitate AI implementation. The 2023 review by van der Vegt et al. was the only review that discussed how the definition of the outcome of sepsis could affect the algorithm’s performance. Depending on which outcome definition was used (e.g. initial inflammatory process vs multi-organ dysfunction) the algorithm could either predict the likelihood of developing sepsis or just diagnose sepsis^16^.

#### Domain 2: The Technology

Lack of robust technological infrastructure emerged as a key impediment to scaling-up AI-based solutions. Outdated networks, limited bandwidth, or poorly maintained servers produced delays in AI output, undermining clinicians’ confidence in the AI solution’s recommendations^2,16–21^. It was recommended across the literature that AI needs to be able to integrate within existing computational resources. Hassan et al. (2023) stressed the significance of embedding AI solutions into existing IT infrastructure with minimal disruption. Kamel Rahimi et al., emphasised that technology fails when it does not fit smoothly into daily workflows – for example using an external dashboard often demanded that staff switch between screens, leading to fragmentation and fatigue^17^.

Concerns around the accuracy of outputs from the AI were also raised across the literature. The potential for false positive warnings led to alert fatigue among users, and inaccurate outputs created burden to the user’s workflow, impacting user perspectives of the AI and impeding adoption^2,16–19,21,22^. Mitigation strategies to address these concerns were raised in the publications and included technical validation, followed by clinical validation. Additional barriers included the concerns around the quality and relevance of the data being fed into AI platforms^2,6,8,17,21,22^, and that AI systems could perpetuate biases present in the data they were trained on^2,6,19,21^. One recurring issue across the studies was the “black-box” phenomenon^2,6,18^. Many advanced AI models lacked interpretability or transparency, making it difficult for clinicians to trust or fully adopt them. Other concerns from potential end users included whether the AI model had been externally validated to assess its generalisability, and whether the outputs produced by the AI could be generalisable across different departments or hospitals^16,17,21^. This domain revealed that transparent, user-friendly, and well-integrated technology is a key prerequisite for the scaling and spread of AI solutions in hospitals.

#### Domain 3: The Value Proposition

The third NASSS domain examined whether the innovation filled a recognised need for diverse stakeholders: patients, clinicians, administrators, and technology developers. Multiple reviews observed that the perception of the value of the AI solution can either catalyse or block widespread adoption.

Kamel Rahimi et al. indicated that end users were more inclined to support the AI solution if it delivered measurable gains through cost savings and improved clinical outcomes^17^. Some examples of gains included reduction in surgery cancelations, reductions in patient’s length of stay in hospitals and improved efficiency of clinician’s workflows^19,23^. Other reviews highlighted that a lack of empirical evidence on the added value of AI application in certain practices could act as a barrier to its implementation^2,6,18,24^. The review by Hassan et al. mentioned that an enabler to AI implementation would be to identify a problem that the AI should address, rather than implementing the AI solution without a use case^2^. Similarly, Kamel Rahimi et al. shared how concerns from end users regarding whether new AI solutions were relevant to their workflow could be a barrier to adoption^17^.

#### Domain 4: The Adopter System

The fourth NASSS domain addressed users’ potential and willingness to adopt the technology—specifically, healthcare professionals, hospital administration teams, data scientists, and patients. The reviews consistently found that staff receptiveness to AI was influenced by factors such as awareness, training, perceived autonomy, and trust.

A lack of trust was frequently reported across the publications due to a misunderstanding of outputs or concerns around the accuracy of the AI outputs^2,6,16,17,21,23,24^. Reviews found that trust improved when users actually had experienced an AI solution making correct predictions^16,21^. Often discussed as a contributor to the scepticism towards AI was the lack of training and education among staff^2,6,7,9,16,18,21,22^. The provision of training for the interpretation of AI outputs, and raising awareness among staff in regard to using AI could help to facilitate trust and uptake in AI implementation^2,16–21,24^.

Some of the reviews identified a fear among clinicians that AI could undermine their professional expertise or lead to job displacement, this finding was similar to reviews that reported that balancing the use of AI with clinician autonomy could be an enabler to its implementation^2,6,16,18,19,24^. Factors cited in the literature to address these concerns included co-designing the AI solutions with those who were going to implement the solution or be affected by its implementation such as patients, healthcare professionals, and data scientists^2,16,17,21,22,24^. The literature also cited the importance of developing a multi-disciplinary team including data scientists and clinicians who would be involved in implementing the AI solution together, to improve team understanding of the solution.

Overall, the adopter system domain underscores that successful AI implementation requires a workforce that sees genuine benefits, feels equipped to use the tool, and experiences no existential threat to professional identity. Without these conditions, scale-up is likely to stall.

#### Domain 5: The Organisation

The fifth domain covered factors that affect an organisation’s ability to adopt AI systems, such as funding decisions, leadership involvement, and readiness to adapt operational processes. Many reviews detailed how organisational culture and resource constraints can impede new technologies^17,18^. The reviews highlighted that the organisational cost for AI implementation could be a barrier to its spread and scalability, specifically the cost for the AI solution, the costs associated with integrating the AI into existing infrastructure, the cost for training staff and the cost for reserving time for staff or appointing new job roles to implement the AI solution^2,6,16–19,21,22^. Another point was raised in relation to sustainability and that there would be costs associated with maintaining the AI and maintaining a level of training among staff.

The synergy between senior leadership commitment and AI expansion emerged as a vital enabler in multiple reviews^2,16,17,21^. Kamel Rahimi et al. highlighted that leadership backing allowed hospital teams to allocate staff time and resources to implement AI solutions. Another structural issue was how AI intersects with existing workflows, the publications cited that a facilitator to AI implementation would be non-disruptive integration or staggered integration to existing workflows^2,16,17,21^. Other facilitators discussed in the reviews included having an implementation plan to provide guidance on the adoption of the AI solution, including aspects such as who would be responsible for receiving the alerts^2,17,21^.

#### Domain 6: The Wider Context

NASSS’s sixth domain addressed political, regulatory, economic, and sociocultural factors. A cross-cutting finding from these reviews was that the external policy environment based on regulatory bodies and government actions can either bolster or undermine local AI initiatives. The authors repeatedly cited that the trajectory of the AI solution in the hospital setting is shaped by external bodies, from government agencies to professional societies and accreditation institutions. Many of the reviews discussed the need for action on: financial and legal endorsement^2,6^; national regulatory standards that consider ethics around consent, data protection, data ownership, legal liability^2,6,17,19^; and an agreed standard for the accuracy of AI before it is approved for clinical practice^2^.

Economic considerations also emerged strongly in the domain of the wider context across the literature. Kamel Rahimi et al. found that financial challenges in health systems could create favourable conditions for AI if the solution was efficient and produced cost saving, but at the same time the high cost of AI could be a barrier for many health systems^17^. Hassan et al. flagged the need for healthcare systems implementing AI solutions to know all the cost upfront from starting, to scaling, and sustaining AI^2^. Ahmed et al. raised the risk of a two-tier health system forming, with some hospitals gaining access to supportive AI solutions whilst others may face barriers due to funding constraints^6^.

#### Domain 7: Interactions Between Domains and Adaptation Over Time

The final NASSS domain focused on how the previous six domains interlinked with each other, over time, shaping the resilience and long-term viability of AI interventions. The included reviews collectively showed that complex interdependencies determine whether an AI solution is implemented. Most commonly intertwined were the domains technology and the adopter system: the complex outputs from AI solutions could lead to a lack of understanding among hospital staff leading to barriers in adopting the technology^17,21,23^. Potential mitigation measures to the lack of understanding included conducting user-feedback and pilots that could lead to iterative updates to the AI solution, making the outputs more user-friendly, or providing more context and transparency behind the technology^2,16–19,21,24^.

## Discussion

This umbrella review has offered a broad perspective on the factors acting as barriers and facilitators influencing the adoption, spread and sustainability of AI implementation in hospital settings. Although the included systematic reviews covered diverse clinical contexts—from operating room management, infection prevention, critical care documentation and wearable patient monitoring, certain cross-cutting insights emerged.

The most common factor that affected adoption was hospital staff perspectives on the trust and understanding of the AI solution^2,6,9,16–24^. Implementation science has long shown that healthcare professionals’ scepticism or poor understanding of a new intervention routinely obstructs roll-out. This is consistent with existing literature on healthcare professionals’ perspectives on e-health interventions, or complex interventions more generally whereby a lack of understanding, lack of training or lack of trust for a specific intervention or solution can affect implementation^25,26^. National and local organisations seem to recognise the importance in ensuring an understanding of AI systems, NHS England have developed a report on developing healthcare workers’ confidence in AI, which sets outs strategies to develop training curricula on using AI^27,28^. Local hospitals sites are following suit: Bedfordshire Hospitals NHS Foundation Trust issued a dedicated Artificial Intelligence Policy in 2024 that mandates staff training on AI fundamentals^29^. Such policies may help to improve the trust and understanding gaps flagged above.

Spread and scalability can be facilitated if AI solutions are externally validated to ensure generalisability across different contexts such as departments and hospitals^16,17,21^. Generalisability of research findings across different hospitals is a goal across most health research^30^. However, there is a growing debate that the generalisability of machine learning models across different hospitals is unachievable due to differences in the operational characteristics of hospitals, instead it may be more appropriate to ensure the algorithm works well within one local setting^31–33^.

To facilitate the sustainability of AI solutions, the cost of maintaining the AI solution in terms of technology and staff training needs to be considered^2,6,16–19,21,22^. Addressing system level factors such as funding and regulations to sustain AI implementation, is an approach that the UK Government is now pushing forward – with funding and plans for supporting AI implementation within the NHS^34,35^.

The findings from this umbrella review can support the future implementation of AI solutions in hospital settings. It is evident from the published studies that AI solutions that are co-produced with clinicians, patients and other end-users, tested in the real world and refined in an iterative way are more likely to respond to the needs of users and be implemented successfully. Furthermore, support from hospital senior leaders can facilitate uptake.

The strengths of this review include that it has been guided by the NASSS framework from the outset, ensuring that our data extraction and synthesis systematically captured the domain insights regarding AI implementation and sustainability in hospitals. This research leveraged a large, multidisciplinary research team, allowing us to conduct double screening at both title and abstract and full-text stages on a subsample of the articles, as well as double data extraction for a subsample of publications, and a full double quality appraisal on all included articles. These measures aimed to minimise bias and enhance the reliability of our findings. Although we used the ROBIS tool for critical appraisal, all reviews were judged to have a high risk of bias, reflecting both the wide heterogeneity of included papers and the fact that ROBIS is more tailored to quantitative systematic reviews. It does, however, remain critical to note that many of the reviews included in this umbrella analysis face their own methodological shortcomings, as indicated in the risk-of-bias assessments. We also recognise that as seven of the included 13 reviews contained some of the same included literature, this may have caused limitations in our research by over-representing themes.

In summary, this application of the NASSS framework shows that implementing AI in hospital settings is a complex undertaking, involving individual, organisational, and technical factors. The emergent conclusion is that sustainable AI adoption requires thoughtful alignment across each NASSS domain.

## Conclusion

This umbrella review pinpointed both the facilitators and barriers for adopting AI solutions in hospitals, and through the NASSS framework we have been able to emphasize the role of technology, value, the adopter system, the organisation and the wider context. The common factors that affected adoption included a lack of trust in and understanding of the AI solutions, whilst scale-up and spread was affected by the generalisability of the AI models. Sustainability was affected by the availability of funds and resources to maintain the AI solution. Some of the factors that acted as facilitators in implementation included co-design activities, comprehensive training, and gaining buy-in from senior leadership. Implementation could also be facilitated by regulatory bodies who can produce transparent governance frameworks and policies that address funding, data security and legal liability.

## Data Availability

All data produced in the present study are available upon reasonable request to the authors

## Appendix

### 1. Appendix 1. Search strategy

**MEDLINE – 13 January 2025**

Ovid MEDLINE(R) ALL <1946 to January 10, 2025>

|  |  |  |
| --- | --- | --- |
| 1 | artificial intelligence.ti,ab. | 54209 |
| 2 | AI.ti,ab. | 64605 |
| 3 | "A.I".ti,ab. | 64605 |
| 4 | machine learning.ti,ab. | 118630 |
| 5 | deep learning.ti,ab. | 66707 |
| 6 | hospital.ti,ab. | 1361586 |
| 7 | tertiary care.ti,ab. | 74189 |
| 8 | secondary care.ti,ab. | 9495 |
| 9 | acute care.ti,ab. | 29675 |
| 10 | inpatient care.ti,ab. | 7983 |
| 11 | intensive care.ti,ab. | 210213 |
| 12 | ward.ti,ab. | 50210 |
| 13 | systematic review.ti,ab. | 330383 |
| 14 | meta-analysis.ti,ab. | 282907 |
| 15 | meta analyses.ti,ab. | 77291 |
| 16 | meta-synthesis.ti,ab. | 1914 |
| 17 | exp Artificial Intelligence/ | 220504 |
| 18 | exp Hospitals/ | 331931 |
| 19 | exp Hospital Units/ | 144383 |
| 20 | exp "Systematic Review"/ | 284913 |
| 21 | exp Meta-Analysis/ | 211143 |
| 22 | 1 or 2 or 3 or 4 or 5 or 17 | 368100 |
| 23 | 6 or 7 or 8 or 9 or 10 or 11 or 12 or 18 or 19 | 1762789 |
| 24 | 13 or 14 or 15 or 16 or 20 or 21 | 528255 |
| 25 | 22 and 23 and 24 | 457 |

### CINAHL Plus – 13 January 2025 (no limits on date of publication or language)

| Search ID | Search Terms | Results |
| --- | --- | --- |
| S1 | AB ("artificial intelligence" OR "ai" OR "a.i." OR "machine learning" OR "deep learning") | 24, 765 |
| S2 | (MH "Artificial Intelligence+") | 38,542 |
| S3 | (MH "Hospitals+") OR (MH "Hospital Units+") | 250,655 |
| S4 | AB ("hospital" OR "tertiary care" OR "secondary care" OR "acute care" OR "inpatient care" OR "intensive care" OR "ward") | 425,059 |
| S5 | (MH "Literature Review+") | 160,750 |
| S6 | AB ("systematic review" OR "meta-analysis" OR "meta-analyses" OR "meta-synthesis") | 122,634 |
| S7 | S1 OR S2 | 52,043 |
| S8 | S3 OR S4 | 583,356 |
| S9 | S5 OR S6 | 216,733 |
| S10 | S7 AND S8 AND S9 | 139 |

### HMIC – 13 January 2025

HMIC Health Management Information Consortium <1979 to November 2024>

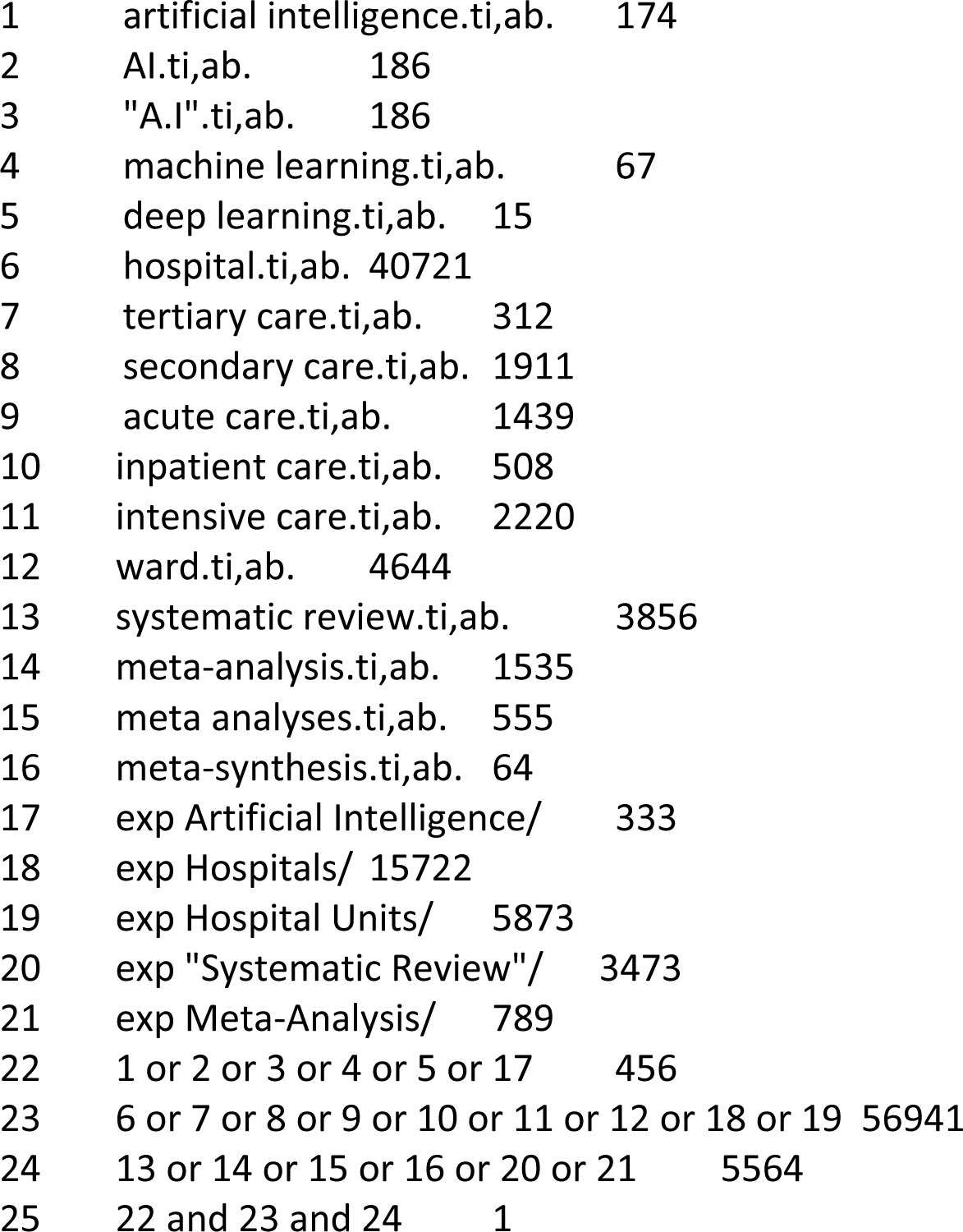

### Cochrane Reviews – 13 January 2025

”artificial intelligence” OR “ai” OR “a.i.” OR “machine learning” OR “deep learning” in Title Abstract Keyword AND “hospital” OR “tertiary care” OR “secondary care” OR “acute care” OR “inpatient care” OR “intensive care” OR “ward” in Title Abstract Keyword AND “systematic review” OR “meta-analysis” OR “meta-analyses” OR “meta-synthesis” in Title Abstract Keyword

Results: 3

### Web of Science – 13 January 2025

”artificial intelligence” OR “ai” OR “a.i.” OR “machine learning” OR “deep learning” in Topic AND “hospital” OR “tertiary care” OR “secondary care” OR “acute care” OR “inpatient care” OR “intensive care” OR “ward” in Topic AND “systematic review” OR “meta-analysis” OR “meta-analyses” OR “meta-synthesis” in Topic

Results: 363

### Appendix 2. List of included articles

- Bellini V, Russo M, Domenichetti T, Panizzi M, Allai S, Bignami EG. Artificial Intelligence in Operating Room Management. *J Med Syst*. 2024;48(1):19. doi:10.1007/s10916-024-02038-2
- Kamel Rahimi A, Pienaar O, Ghadimi M, et al. Implementing AI in Hospitals to Achieve a Learning Health System: Systematic Review of Current Enablers and Barriers. *J Med Internet Res*. 2024;26:e49655. doi:10.2196/49655
- Kucukkaya A, Arikan E, Goktas P. Unlocking ChatGPT’s potential and challenges in intensive care nursing education and practice: A systematic review with narrative synthesis. *Nurs Outlook*. 2024;72(6):102287. doi:10.1016/j.outlook.2024.102287
- Lee C, Britto S, Diwan K. Evaluating the Impact of Artificial Intelligence (AI) on Clinical Documentation Efficiency and Accuracy Across Clinical Settings: A Scoping Review. *Cureus*. Published online November 19, 2024. doi:10.7759/cureus.73994
- Piaggio D, Zarro M, Pagliara S, et al. The use of smart environments and robots for infection prevention control: A systematic literature review. *Am J Infect Control*. 2023;51(10):1175-1181. doi:10.1016/j.ajic.2023.03.005
- Baig MM, GholamHosseini H, Moqeem AA, Mirza F, Lindén M. A Systematic Review of Wearable Patient Monitoring Systems – Current Challenges and Opportunities for Clinical Adoption. *J Med Syst*. 2017;41(7):115. doi:10.1007/s10916-017-0760-1
- Tan S, Mills G. Designing Chinese hospital emergency departments to leverage artificial intelligence—a systematic literature review on the challenges and opportunities. *Front Med Technol*. 2024;6. doi:10.3389/fmedt.2024.1307625
- van der Vegt AH, Campbell V, Mitchell I, et al. Systematic review and longitudinal analysis of implementing Artificial Intelligence to predict clinical deterioration in adult hospitals: what is known and what remains uncertain. *Journal of the American Medical Informatics Association*. 2024;31(2):509-524. doi:10.1093/jamia/ocad220
- van der Vegt AH, Scott IA, Dermawan K, Schnetler RJ, Kalke VR, Lane PJ. Deployment of machine learning algorithms to predict sepsis: systematic review and application of the SALIENT clinical AI implementation framework. *Journal of the American Medical Informatics Association*. 2023;30(7):1349-1361. doi:10.1093/jamia/ocad075
- Hassan M, Kushniruk A, Borycki E. Barriers to and Facilitators of Artificial Intelligence Adoption in Health Care: Scoping Review. *JMIR Hum Factors*. 2024;11:e48633. doi:10.2196/48633
- Ahmed MI, Spooner B, Isherwood J, Lane M, Orrock E, Dennison A. A Systematic Review of the Barriers to the Implementation of Artificial Intelligence in Healthcare. *Cureus*. Published online October 4, 2023. doi:10.7759/cureus.46454
- Lambert SI, Madi M, Sopka S, et al. An integrative review on the acceptance of artificial intelligence among healthcare professionals in hospitals. *NPJ Digit Med*. 2023;6(1):111. doi:10.1038/s41746-023-00852-5
- Ouanes K, Farhah N. Effectiveness of Artificial Intelligence (AI) in Clinical Decision Support Systems and Care Delivery. *J Med Syst*. 2024;48(1):74. doi:10.1007/s10916-024-02098-4

